# The effect of mobile applications for menstrual cycle or fertility trackers on women’s health: protocol for a systematic review

**DOI:** 10.1101/2020.09.25.20201251

**Authors:** Andrea I. Nomura-Sakata, Cinthya I. Mogrovejo-Navas, Alejandra Navarro-Grau, Jessica Hanae Zafra-Tanaka

**Affiliations:** School of Medicine, Universidad Peruana Cayetano Heredia, Lima, Perú; CRONICAS, Centro de Excelencia En Enfermedades Crónicas, Universidad Peruana Cayetano Heredia, Lima, Perú

**Author notes:** Correspondence Alejandra Navarro Grau Delgado.

**Keywords:** Menstrual cycle, mobile application, tracking, women’s health

## Abstract

**Background:** Nowadays the use of smartphones and the development of health-related mobile applications has increased worldwide. Menstrual cycle tracking applications (MCTAs) have become especially popular among women because of their practicality in recording menstrual cycles, characteristics of bleeding and prediction of cycle stages. There are various studies regarding the use of MCTAs for different aspects of women’s health such as estimating a fertility window for both conception and contraception, help register last menstrual period for calculation of gestational age, record pre-menstrual symptoms, among others. However, effects of MCTAs have not been analyzed in a systematic review.

**Objective:** The aim of this study is to evaluate the effect of mobile applications for menstrual cycle or fertility trackers on women’s health.

**Methods:** A systematic review will be conducted, starting with a search in PubMed, CENTRAL and Scopus using search terms related to mobile applications and menstrual or fertility tracking. Only randomized controlled trials will be screened with a sample of child-bearing aged women that use menstrual or fertility tracking mobile applications. Selected studies will be fully analyzed and the results will be recorded on a spread sheet. Study selection and data extraction will be conducted by two reviewers independently and the Cochrane Risk of Bias Tool for RCT will be used for assessment of risk of bias. Discrepancies will be reviewed with a third reviewer.

**Conclusion:** Currently, there is a lack of information on the effects of using MCTAs on women’s health. This systematic review aims to provide an analysis on the outcomes of the usage of these applications and evaluate any potential effects.

**Conflicts of interest:** All authors declare to have no conflicts of interest.

## Background

Since the introduction of smartphones, their availability, practicality and wide range of uses has increased exponentially worldwide as well as the development of health-related mobile applications. By 2018, 75% of women in the USA own and use a smartphone and of those, 94% of women are within a child-bearing age^1^.Menstrual cycle tracking applications (MCTAs) have become increasingly popular and are currently among the most popular health apps, especially in adolescent women^2^.

MCTAs record length of menstrual cycles, volume and duration of bleeding, predict menstrual cycle stages and help users follow menstrual related symptoms. This is especially important since symptoms affect mental and physical health of women as well as their productivity in a work environment which is why correlating symptoms with a specific stage of the cycle can help alleviate or reduce them^3^. Having an accurate record of menstruation dates is beneficial in estimating gestational age and estimated birth dates in pregnant women. In low resource areas where ultrasound is not readily available, the date of the last menstrual period is essential; thus, relying on an app could be more precise than memory^4^. Also, these apps have proven to aid in the identification of the fertility window for couples wishing to conceive and helpful regarding contraception and avoiding unintended pregnancies by estimating the next period date^5^.

Previous systematic reviews regarding MCTAs have focused on the features and quality of the applications, but studies investigating the outcomes and effects of using MCTAs have not yet been published. Considering the popularity and the importance given to the MCTAs by the users would be valuable to analyze their possible benefits. The aim of this study is to evaluate the effect of mobile applications for menstrual cycle or fertility trackers on women’s health.

## Methods

### Study design

This systematic review will be conducted according to the preferred reporting items for systematic reviews and meta analyses guidelines (PRISMA)^6^.

### Eligibility criteria

1. **Participants**: This study will include child-bearing aged women without restriction on location
2. **Intervention**: Use of mobile applications for menstrual cycle or fertility tracking
3. **Comparison**: Any control group, including no tracking, paper tracking or any other.
4. **Types of studies**: This study will include randomized controlled trials with no publication date restriction

### Exclusion criteria

1. The following study designs: cross-sectional studies, case control, cohort, case reports, case series, letters to the editor, editorial, narrative review, systematic reviews, correspondence, short communications, technical notes, commentaries and pictorial essays.
2. Studies that evaluate the quality of the application and no health-related outcome is mentioned
3. Studies that include applications that are not related to the menstrual cycle or fertility tracking such as: information providing-only or applications used to complement a medical appointment like ones used in waiting rooms
4. Studies published in any language other than Spanish or English

### Literature search, data collection and coding

- The following databases will be searched: 1) PubMed, 2) Central Cochrane Library, and 3) Scopus.
- The references of included studies will also be reviewed to find more potentially eligible studies.
- The complete search strategy for each database, the number of hits retrieved, and the reasons for exclusion of any studies at the full text stage of selection will be recorded and provided as appendices. An overview of the search strategy can be found in **Appendix 1.**
- Two authors will independently screen titles and abstracts looking for potential studies to be included and proceed to read in detail the full text of the selected studies following the inclusion and exclusion criteria. Any conflict will be resolved by a discussion to achieve consensus.
- The same three authors will record the results of included and excluded publications and show them on a PRISMA flow diagram.
- Duplicate articles will be removed using a spreadsheet (Microsoft Excel Software)
- The data extracted from the selected articles will include:
  ○ Study details: first author, corresponding author, article title, country, year of publication and year of data collection
  ○ Subjects: number of participants, range of age, population source, location, inclusion and exclusion criteria of studies selected
  ○ Study methodology: name of the tool, type of information recording, frequency of recording, among others.
  ○ Results: outcome of study

In case we find a study in which the methodology or outcome is not clearly specified, we will try to contact the corresponding author. If we do not receive an answer, then the study will be excluded from the systematic review.

### Risk of bias (quality) assessment

We will assess risk of bias using the Cochrane Risk of Bias Tool for RCT^7^ and present a table with the results of this assessment.

## Statistical analysis

We will collect information on the effects of outcomes assessed in the primary studies and conduct a meta-analysis to estimate pooled-effects as standardized or weighted mean difference or relative risk (RR) with corresponding 95% confidence intervals for continuous or dichotomous data, respectively.

## Data Availability

All relevant information will be published as supplementary materials

